# Predictors of Incident Viral Symptoms Ascertained in the Era of Covid-19

**DOI:** 10.1101/2020.09.24.20197632

**Authors:** Gregory M Marcus, Jeffrey E Olgin, Noah D Peyser, Eric Vittinghoff, Vivian Yang, Sean Joyce, Robert Avram, Geoffrey H Tison, David Wen, Xochitl Butcher, Helena Eitel, Mark J Pletcher

## Abstract

**Background:** In the absence of universal testing, effective therapies, or vaccines, identifying risk factors for viral infection, particularly readily modifiable exposures and behaviors, is required to identify effective strategies against viral infection and transmission.

**Methods:** We conducted a world-wide mobile application-based prospective cohort study available to English speaking adults with a smartphone. We collected self-reported characteristics, exposures, and behaviors, as well as smartphone-based geolocation data. Our main outcome was incident symptoms of viral infection, defined as fevers and chills plus one other symptom previously shown to occur with SARS-CoV-2 infection, determined by daily surveys.

**Findings:** Among 14, 335 participants residing in all 50 US states and 93 different countries followed for a median 21 days (IQR 10-26 days), 424 (3%) developed incident viral symptoms. In pooled multivariable logistic regression models, female biological sex (odds ration [OR] 1.75, 95% CI 1.39-2.20, p<0.001), anemia (OR 1.45, 95% CI 1.16-1.81, p=0.001), hypertension (OR 1.35, 95% CI 1.08-1.68, p=0.007), cigarette smoking in the last 30 days (OR 1.86, 95% CI 1.35-2.55, p<0.001), any viral symptoms among household members 6-12 days prior (OR 2.06, 95% CI 1.67-2.55, p<0.001), and the maximum number of individuals the participant interacted with within 6 feet in the past 6-12 days (OR 1.15, 95% CI 1.06-1.25, p<0.001) were each associated with a higher risk of developing viral symptoms. Conversely, a higher subjective social status (OR 0.87, 95% CI 0.83-0.93, p<0.001), at least weekly exercise (OR 0.57, 95% CI 0.47-0.70, p<0.001), and sanitizing one’s phone (OR 0.79, 95% CI 0.63-0.99, p=0.037) were each associated with a lower risk of developing viral symptoms.

**Interpretation:** While several immutable characteristics were associated with the risk of developing viral symptoms, multiple immediately modifiable exposures and habits that influence risk were also observed, potentially identifying readily accessible strategies to mitigate risk in the Covid-19 era.

**Funding:** This study was funded by IU2CEB021881-01 and 3U2CEB021881-05S1 from the NIH/ NIBIB to Drs. Marcus, Olgin, and Pletcher.

**Research in context:** *Evidence before this study:* Predictors of incident viral infection have been determined largely from cross-sectional studies prone to recall bias among individuals representing geographically constrained regions, and most were conducted before the era of the current Covid-19 pandemic.

*Added value of this study:* We conducted a world-wide, mobile application-based, longitudinal cohort study utilizing time-updated predictors and outcomes, providing novel and current information regarding risk-factors for incident viral symptoms based on real-time information in the era of Covid-19.

*Implications of all the available evidence:* These data suggest that certain immutable characteristics influence the risk for incident viral symptoms, while smoking cessation, physical distancing to avoid contact with individuals outside the household, regular exercise, and sanitizing one’s phone may each help mitigate the risk of viral infection.

## Introduction

The global SARS-CoV-2 pandemic has affected communities in every habitable continent and every state in the US. Given what is generally known about respiratory viruses, strategies to mitigate transmission have included government orders to practice regular hand hygiene, physical distancing, including the closures of public locations commonly associated with community gatherings, and more recently to wear masks.^1-3^ Studies thus far have largely focused on comparisons among those seeking medical care for the disease^4-7^ or evaluations of large administrative datasets.^8^ It can be difficult to track individual-level characteristics and behaviors, particularly as they are dynamic and changing over time, as they relate to incident disease. Members of the public may benefit by understanding strategies under their direct control that may influence their own risk of infection and viral transmission.

Tracking viral infection is hindered by the absence of universal and repeated testing. In the absence of such testing, recent evidence suggests that symptoms themselves may be useful markers of SARS-CoV-2 infection.^9^ While the virus may be asymptomatic, a variety of symptom clusters associated with the disease have been identified, often including fever, but ranging from typical respiratory symptoms to gastrointestinal afflictions to somewhat idiosyncratic findings such as anosmia/ageusia and conjunctivitis.^10-18^ In the past, ascertainment of viral symptoms has relied on assessments of those seeking medical care or retrospective surveys that may be prone to recall bias. Given the current near-ubiquity of smartphones and use of related mobile apps, technology is now available to regularly and repeatedly query large numbers of individuals over time, providing access to symptom development as it arises. Although monitoring for viral symptoms may be neither sufficiently sensitive nor specific for SARS-CoV-2 infection, these outcomes are, by their nature, inherently experienced by the individual, potentially providing valuable information that may be best leveraged by modern mobile technology.

We sought to use prospectively collected information about exposures and modifiable behaviors, along with daily symptom reporting, to identify risk factors for incident viral symptoms using a globally-available, smartphone mobile application-based study, the COVID-19 Citizen Science Study.

## Methods

We launched the COVID-19 Citizen Science Study, a mobile application-based study compatible with Android or iOS operating systems, on March 26, 2020. The mobile application was built by investigators and developers at the University of California, San Francisco, using the NIH-supported Eureka digital and mobile research platform. Enrollment is open to any adult with a smartphone, and study information has been broadcast via press release, social media, and to participants in the Eureka-based Health eHeart Study. Participants were encouraged to recruit additional individuals. Updated study information, including number of participants, maps of symptom clusters, and location of participants around the world can be found at https://covid19.eurekaplatform.org/. All participation is remote, without geographic restriction. Verification of cell phone numbers via text was required before proceeding from study registration to remote-based study consent and subsequent study participation. Retention strategies include daily notifications, data visualizations, and intermittent study update blog posts. Our Citizen Scientist participants contribute study question ideas that are then included into the study and reported to participants as participant-generated.

Participants complete surveys written in lay language meeting Flesch-Kincaid criteria for an 8^th^ grade reading level (https://readabilityformulas.com). At baseline, surveys collected information about demographics, education, occupation, SARS-CoV-2 status (referred to in surveys as “the novel coronavirus, the virus that causes COVID-19”), behaviors, living conditions, attitudes regarding the SARS-CoV-2 pandemic, local government restrictions related to the disease, medical conditions, and medications (the surveys are included in the ***Supplementary Appendix***). Perceived socioeconomic status was assessed using the MacArthur subjective social status ladder,^19,20^. All participants received an optional invitation to share their smartphone-based geolocation data.

Participants receive a daily survey, timed to occur synchronously to their local same time of day when they engaged with the first baseline survey, via mobile application-based push notification. The daily survey includes queries about current viral symptoms, updated according to new information, using “check all that apply” including: “A scratchy throat”; “A cough (worse than usual if you have a baseline cough)”; “A painful sore throat”; “A temperature greater than 100·4 °F or 38·0 °C”; “A runny nose”; “Symptoms of fever or chills”; “Muscle aches (worse than usual if you have baseline muscle aches)”; “Shortness of breath”; “Nausea, vomiting or diarrhea” (added March 30, 2020); “Unable to taste or smell” (added March 31, 2020); “Red or painful eyes” (added April 13, 2020); or “none of the above.” The daily survey then includes questions regarding current symptoms among household contacts and the number of individuals outside the household the participant interacted with within six feet (about 1·83 meters) in the previous 24 hours.

Participants received weekly surveys to update information regarding sleep, exercise, hand hygiene, social and physical distancing behaviors, habits such alcohol consumption, and SARS-CoV-2 infection status. All surveys remained open for 24 hours.

For those that consented to geolocation tracking, smartphone-based geolocation using a combination of the Global Positioning System and cell phone tower triangulation was collected every 5 minutes for Android phones and whenever the phone accelerometer exhibited movement (in order to minimize battery drain) for iOS smartphones. Geolocation latitude and longitude coordinates were clustered within an individual for every day of the study using the HDBScan clustering algorithm. The most prevalent cluster of geolocation coordinates for a user was defined as “home.” Time spent at a cluster was calculated as the time difference between the current location cluster and any future cluster change. Daily time spent at home was calculated as the time spent at the cluster identified as “home” divided by the time between the first and last coordinate collected daily. In addition, distance travelled was calculated as the sum of the successive distances between all consecutive coordinates collected within a user on a daily basis. Long-distance travel was defined as movement of at least 1,000 kilometers within 24 hours.

Occupation was dichotomized into healthcare workers versus not; sleep was determined as the average number of hours per day over each week; exercise, defined as physical activity for at least 20 minutes that resulted in breathing heavily or to “break a sweat,” was dichotomized into more or less than once weekly; alcohol was assessed as average daily standard drinks; cigarette, e-cigarette, and marijuana use were dichotomized into any use in the last 30 days versus not; household symptoms were dichotomized into any versus none in the previous 6-12 days; and the maximum number of contacts within six feet (about 1·83 meters) reported in the previous 6-12 days were derived from the daily survey responses.

For the current analyses, all participants reporting a previous positive test for SARS-CoV-2 and those with any symptoms upon entry to the study were excluded. Those with baseline medical conditions that might themselves contribute to the symptoms of interest, including atrial fibrillation, coronary artery disease, congestive heart failure, chronic obstructive pulmonary disease, and asthma, were excluded. Based on the daily surveys, incident viral symptoms were defined as the first report of a combination of fever or chills plus at least one other symptom on the same day. Follow-up for the current study ended May 3, 2020. The study was approved by the University of California, San Francisco Institutional Review Board. All participants provided informed electronic consent.

### Statistical Analyses

Normally distributed continuous variables are presented as means ± SD and compared using t-tests, where continuous variables with skewed distributions are presented as medians with interquartile ranges (IQR) and compared using Wilcoxon rank sum tests. Categorical variables were compared using chi-squared tests. Pooled logistic regression models were used to identify factors associated with incident symptoms, potentially including baseline characteristics (demographics, medical conditions, habits, and behaviors related to viral infection risk such as hand hygiene), and time-updated information from daily and weekly surveys. Exposures that expected to influence the risk of viral infection that would then manifest as future symptoms several days later were evaluated using survey data for 6-12 days earlier. Consequently, only participants with at least one daily survey at least 6 days after the first were included in the pooled logistic regression models. Beginning with the subset of variables associated with incident symptoms at p < 0·1 in pooled logistic regression models adjusting only for age, sex, race, and calendar date (with linear and non-linear components), backward deletion was used to select multivariable models retaining covariates with p < 0·05. Statistical analyses were performed using Stata, version 16 (College Station, TX). Two-tailed p-values < 0·05 were considered statistically significant.

### Role of the Funding Source

The funder, NIH/ NIBIB, did not influence the concept, creation, or conduct of the study.

## Results

After exclusions were applied, 14,335 participants were available and contributed to the incident analyses. Differences between these participants and those that entered the study reporting at least one viral symptom are shown in **Table 1**. Participants resided in all 50 states and in 93 countries outside the US. While a mean 42% ± 12% of all participants completed the daily survey each day, 95%-100% of all participants completed at least one daily survey per week throughout the study period, and weekly surveys were completed 66 ± 26% of the time (***Supplementary Table 1***).

**Table 1.** Baseline characteristics of participants with and without prevalent viral symptoms.

|  | **Prevalent Symptoms**<br>**N=374** | **Included in Incident**<br>**Analyses**<br>**N=14,335** | **p-value** |
| --- | --- | --- | --- |
| **Age Category** |  |  | <0.001 |
| 18-29 | 63 (16.8%) | 1,961 (13.7%) |  |
| 30-39 | 112 (29.9%) | 3,225 (22.5%) |  |
| 40-49 | 86 (23.0%) | 2,873 (20.0%) |  |
| 50-59 | 55 (14.7%) | 2,839 (19.8%) |  |
| 60+ | 58 (15.5%) | 3,437 (24.0%) |  |
| **Female Biological Sex** | 253 (67.6%) | 9,322 (65.0%) | <0.001 |
| **Race/Ethnicity** |  |  | 0.005 |
| White | 290 (77.5%) | 11,342 (79.1%) |  |
| Black | 7 (1.9%) | 164 (1.1%) |  |
| Hispanic (any race) | 43 (11.5%) | 1,307 (9.1%) |  |
| Asian or Pacific Islander | 17 (4.5%) | 1,150 (8.0%) |  |
| Other (including<br>multiracial) | 17 (4.5%) | 372 (2.6%) |  |
| **Highest Level of**<br>**Education** |  |  | <0.001 |
| Less than high school | 4 (1.1%) | 57 (0.4%) |  |
| High school graduate | 14 (3.7%) | 471 (3.3%) |  |
| Some college | 80 (21.4%) | 2,034 (14.2%) |  |
| College graduate | 119 (31.8%) | 5,039 (35.2%) |  |
| Post-graduate | 149 (39.8%) | 6,566 (45.8%) |  |
| Other | 8 (2.1%) | 167 (1.2%) |  |
| **MacArthur Subjective**<br>**Social Status Ladder** | 7.0 (5.0-8.0) | 7.0 (6.0-8.0) | <0.001 |
| **Health Care Worker** | 73 (19.5%) | 3,044 (21.2%) | 0.42 |
| **Anemia** | 78 (20.9%) | 1,440 (10.0%) | <0.001 |
| **Cancer** | 24 (6.4%) | 558 (3.9%) | 0.044 |
| **Diabetes** | 26 (7.0%) | 701 (4.9%) | 0.18 |
| **High Blood Pressure** | 90 (24.1%) | 3,128 (21.8%) | 0.37 |
| **HIV** | 2 (0.5%) | 77 (0.5%) | 0.44 |
| **Immunodeficiency** | 23 (6.1%) | 322 (2.2%) | <0.001 |
| **Children home from**<br>**college living with you** | 20 (5.3%) | 1,098 (7.7%) | 0.096 |
| **School-age children**<br>**living with you** | 103 (27.5%) | 3,575 (24.9%) | 0.51 |
| **At least weekly exercise** | 240 (64.3%) | 10,627 (74.2%) | <0.001 |
| **Median drinks per day**<br>**(IQR)** | 0.2 (0.0-0.9) | 0.4 (0.0-1.0) | 0.002 |
| **Median hours of sleep**<br>**(IQR)** | 5.0 (5.0-5.0) | 7.0 (7.0-8.0) | 0.076 |
| **Cigarettes: any use in**<br>**last 30 days** | 30 (8.1%) | 703 (4.9%) | 0.003 |
| **E-cigarettes: any use in**<br>**last 30 days** | 24 (6.4%) | 342 (2.4%) | <0.001 |
| **Marijuana: any use in**<br>**last 30 days** | 42 (11.4%) | 1,448 (10.2%) | 0.17 |
| **Any pets at home** | 223 (59.6%) | 8,512 (59.4%) | 0.040 |
| **Received flu shot within the year** | 251 (67.1%) | 10,772 (75.1%) | 0.002 |

Over a median follow-up of 21 days (IQR 10-26 days), 424 (3%) participants developed incident viral symptoms. ***Supplementary Table 2*** shows the specific symptoms reported. **Figure 1** illustrates the locations of participants with and without symptoms. **Figure 2** provides a sample summary of enrollment, survey completion, symptom development, and follow-up over time. In minimally adjusted logistic models adjusting only for age, sex, race, ethnicity, and date, a higher level of education and subjective social scale, exercising at least once weekly, a longer average sleep duration, and sanitizing one’s phone were each associated with a lower risk while a history of anemia, hypertension or some immunodeficiency, cigarette smoking, e-cigarette use, marijuana use, having pets at home, having household members with viral symptoms, and the number of individuals with which the participant interacted with within six feet (about 1·83 meters) each predicted a higher risk of incident viral symptoms (**Table 2**). Pertinent characteristics that failed to exhibit statistically significant relationships included HIV status, hand washing practices, reported government restrictions, and, per geolocation measurements, amount of time at home and daily distance traveled. In the backwards stepwise logistic model, the following were retained: a higher level of subjective social status, exercise, and sanitizing one’s phone were each associated with a significantly lower risk of developing viral symptoms, whereas female sex, a history of anemia, hypertension, recent cigarette smoking, recent household contacts with viral symptoms, and the maximum number of individuals recently in contact with the participant within six feet (about 1·83 meters) were each associated with a significantly heightened risk of developing viral symptoms (**Table 3**).

**Table 2.** Minimally adjusted odds of incident symptoms.

| Characteristic | Odds Ratio | 95% CI | p-value | Group p-value |
| --- | --- | --- | --- | --- |
| **Highest level of education** |  |  |  |  |
| Less than high school | reference |  |  | <0.001* |
| High school graduate | 0.44 | 0.16, 1.16 | 0.10 | <0.001† |
| Some college | 0.51 | 0.21, 1.24 | 0.14 | 0.015# |
| College grad | 0.29 | 0.12, 0.69 | 0.005 |  |
| Post-grad | 0.26 | 0.11, 0.63 | 0.003 |  |
| Other | 0.32 | 0.09, 1.08 | 0.07 |  |
| **MacArthur Subjective Social Status Ladder** | 0.82 | 0.77, 0.86 | <0.001 |  |
| **Health care worker** | 1.01 | 0.81, 1.28 | 0.90 |  |
| **Anemia** | 1.63 | 1.31, 2.03 | <0.001 |  |
| **Cancer** | 1.02 | 0.61, 1.69 | 0.94 |  |
| **Diabetes** | 1.60 | 1.14, 2.26 | 0.007 |  |
| **High blood pressure** | 1.61 | 1.28, 2.02 | <0.001 |  |
| **HIV** | 1.03 | 0.47, 2.25 | 0.94 |  |
| **Immunodeficiency** | 1.58 | 1.26, 1.97 | <0.001 |  |
| **Children home from college living with you** | 1.11 | 0.78, 1.57 | 0.57 |  |
| **School-age children living with you** | 1.20 | 0.98, 1.47 | 0.08 |  |
| **At least weekly exercise** | 0.48 | 0.39, 0.58 | <0.001 |  |
| **Average drinks per day** | 0.90 | 0.76, 1.07 | 0.25 |  |
| **Average sleep duration (hours)** | 0.84 | 0.76, 0.93 | <0.001 |  |
| **Cigarettes: any use in last 30 days** | 2.43 | 1.79, 3.30 | <0.001 |  |
| **E-cigarettes: any use in last 30 days** | 2.00 | 1.41, 2.84 | <0.001 |  |
| **Marijuana: any use in last 30 days** | 1.32 | 1.01, 1.74 | 0.045 |  |
| **Any pets at home** | 1.31 | 1.07, 1.62 | 0.01 |  |
| **Hand washing practices** |  |  |  |  |
| Much more | reference |  |  | 0.54* |
| Somewhat more | 1.17 | 0.94, 1.46 | 0.16 | 0.87† |
| A little more | 1.05 | 0.71, 1.57 | 0.80 | 0.52# |
| No change | 1.21 | 0.62, 2.37 | 0.57 |  |
| Somewhat less | n/a | n/a, n/a | n/a |  |
| Much less | n/a | n/a, n/a | n/a |  |
| **Flu shot** | 0.90 | 0.72, 1.13 | 0.35 |  |
| **Sanitized phone** | 0.73 | 0.58, 0.92 | 0.007 |  |
| **Local government ordinances** |  |  |  |  |
| School closures | 0.76 | 0.57, 1.03 | 0.07 |  |
| Restricted gatherings by venue | 0.84 | 0.57, 1.23 | 0.37 |  |
| Restricted gatherings by number | 1.06 | 0.71, 1.56 | 0.79 |  |
| Recommended working from home | 0·83 | 0·58, 1·18 | 0·29 |  |
| Shelter in place | 0·89 | 0·66, 1·20 | 0·44 |  |
| Other restrictions | 0·95 | 0·77, 1·16 | 0·59 |  |
| **Any household symptoms, 6-12 days ago** | 2·07 | 1·68, 2·56 | <0·001 |  |
| **Maximum contacts (per 10), 6-12 days ago** | 1·21 | 1·12, 1·31 | <0·001 |  |
| **Distance traveled, 6-12 days ago** |  |  |  |  |
| 0 | reference |  |  | 0·29* |
| <=1 km | 0·86 | 0·60, 1·24 | 0·43 | 0·22† |
| >1-10 km | 0·87 | 0·58, 1·29 | 0·49 | 0·15# |
| >10-50 km | 1·07 | 0·70, 1·63 | 0·75 |  |
| >50 km | 0·52 | 0·25, 1·08 | 0·08 |  |
| **Any travel >1000km, 6-12 days ago** | 0·36 | 0·09, 1·47 | 0·15 |  |
| **Time at home, 6-12 days ago** |  |  |  |  |
| <25% | reference |  |  | 0·64* |
| 25-50% | 1·07 | 0·77, 1·50 | 0·69 | 0·55† |
| 50-75% | 0·91 | 0·60, 1·39 | 0·66 | 0·51# |
| 75-100% | 1·18 | 0·88, 1·58 | 0·27 |  |

Models were adjusted for age, sex, race, ethnicity, and date
\* overall heterogeneity

**Table 3.** Independent Predictors of Incident Symptoms. Derived backwards stepwise elimination of covariates (see methods). * overall heterogeneity † heterogeneity of non-reference levels # linear trend

| Characteristic | Odds ratio | 95% CI | p-value | Group p-value |
| --- | --- | --- | --- | --- |
| **Age category** |  |  |  |  |
| 18-29 | reference |  |  | 0.22* |
| 30-39 | 0.83 | 0.60, 1.16 | 0.28 | 0.21† |
| 40-49 | 0.87 | 0.63, 1.22 | 0.42 | 0.15# |
| 50-59 | 0.96 | 0.69, 1.34 | 0.81 |  |
| 60+ | 0.69 | 0.48, 1.00 | 0.049 |  |
| **Race/ethnicity** |  |  |  |  |
| White | reference |  |  | 0.41* |
| Black | 1.04 | 0.46, 2.37 | 0.92 | 0.26† |
| Hispanic (any race) | 1.10 | 0.80, 1.50 | 0.55 | 0.80# |
| Asian or Pacific Islander | 0.73 | 0.48, 1.10 | 0.14 |  |
| Other (including multiracial) | 1.31 | 0.79, 2.16 | 0.30 |  |
| **Female Biological Sex** | 1.75 | 1.39, 2.20 | <0.001 |  |
| **MacArthur Subjective Social Status Ladder** | 0.87 | 0.83, 0.93 | <0.001 |  |
| **Anemia** | 1.45 | 1.16, 1.81 | 0.001 |  |
| **High blood pressure** | 1.35 | 1.08, 1.68 | 0.007 |  |
| **At least weekly exercise** | 0.57 | 0.47, 0.70 | <0.001 |  |
| **Cigarettes: any use in last 30 days** | 1.86 | 1.35, 2.55 | <0.001 |  |
| **Sanitized phone** | 0.79 | 0.63, 0.99 | 0.037 |  |
| **Any household symptoms, 6-12 days ago** | 2.06 | 1.67, 2.55 | <0.001 |  |
| **Maximum contacts (per 10), 6-12 days ago** | 1.15 | 1.06, 1.25 | <0.001 |  |
| **Calendar date (linear)** | 0.93 | 0.90, 0.97 | <0.001 |  |
| **Calendar date (non-linear)** | 1.05 | 1.01, 1.09 | 0.019 |  |

**Figure 1.**
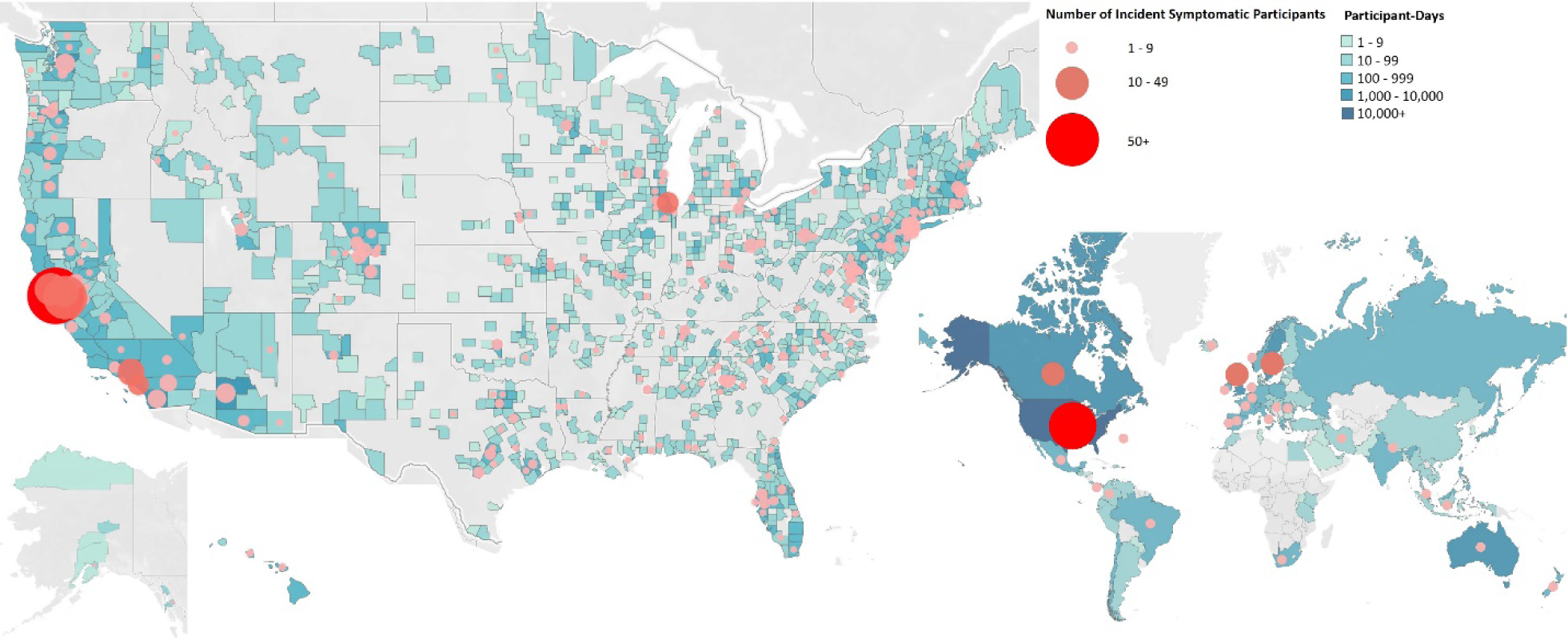
Location of study participants. Blue shading represents gradations of the number of participant-days within the US by county (left) and in the world by nation (right). Red shading depicts the number of symptomatic participants by location.

**Figure 2.**
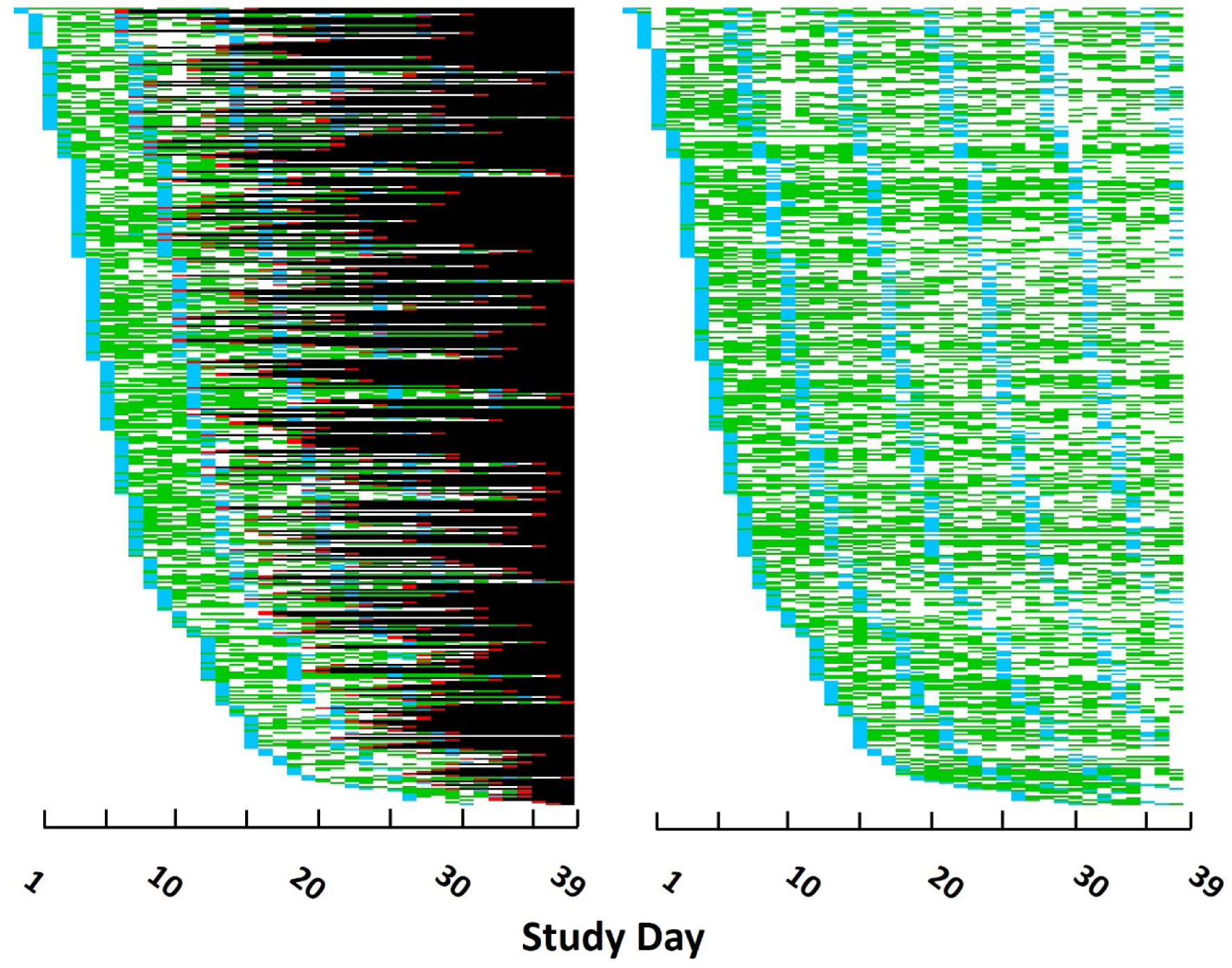
Heat maps of symptomatic and a sample of asymptomatic patients displaying time of enrollment, survey completion, time of symptom development, and follow-up. The left plot depicts participants that developed symptoms. The right plot depicts participants who did not develop symptoms matched in a one-to-one fashion with each symptomatic case by time of enrollment. Each row represents a unique study participant (n=424 for each plot). The X-axis represents days of the current study. Blue=weekly survey completed (the first blue represents the enrollment visit) Green=daily survey completed (the daily survey contents are included in the weekly survey) Red=symptoms developed Black=after development of symptoms White=no data entry prior to or in the absence of symptoms

## Discussion

Among an international cohort involving collection of prospective and time-updated data, female sex, anemia, hypertension, recent cigarette smoking, living with someone with viral symptoms, and the maximum number of recent contacts within six feet (about 1·83 meters) outside the home each predicted a higher risk of developing viral symptoms. Conversely, a higher self-perceived social status, regular exercise, and sanitizing one’s phone were each associated with a lower risk of subsequently reporting viral symptoms.

As of July 25, 2020, there were more than 15 million confirmed cases of SARS-CoV-2 infections and more than 640,000 Covid-19-related deaths around the world.^21^ In response, tremendous investment and efforts are being dedicated to enhance the availability of testing,^22^ identify effective therapies,^23^ and ultimately to develop a vaccine.^24^ Although these remedies are being pursued at an unprecedented pace, the number of infections and deaths continues to grow, and, even after these new technologies, drugs, and vaccines are developed, additional time will be required to disseminate them. While studies of hospitalized patients are valuable, ultimately the characteristics, behaviors, and exposures of individuals in the general community associated with the development of viral symptoms can be helpful in several ways: to identify those at highest risk of developing these symptoms, which may help prioritize protecting the most vulnerable; to provide novel insights regarding the biology of current viral disease; and ideally to identify low-risk and modifiable behaviors that individuals might practice or avoid to reduce their own individual-level risk.

Clearly, self-reported symptoms of a viral infection do not equal SARS-CoV-2 infection. However, the specific viral symptoms repeatedly queried in our population were developed based on available evidence regarding the nature of SARS-CoV-2 infection,^10–18^ and similar viral diseases, including the common cold and influenza for example, very likely share common properties related to an individual’s susceptibility to infection.^25^ In addition, independent of commonalities across easily transmissible viral diseases, there are shared phenomena related to the human immune system’s general vulnerability to viral infection.^26^ Finally, due to the lack of universal testing, there is some evidence that surveillance for viral symptoms may itself have several advantages, often reflecting, either directly or indirectly, underlying Covid-19 disease.^9,27,28^

The higher incidence of viral symptoms among women in our cohort runs counter to the prevailing evidence that men are at higher risk of both SARS-CoV-2 infection and related morbidity and mortality.^29^ While this may suggest the viral symptoms detected by the current study fail to capture patterns most relevant to SARS-CoV-2, these differences may have occurred because our models adjusted for related mediators (such as smoking)^30^ or because women are either more likely to experience or report more mild symptoms. Previous community-based studies often adjust or weight for sex distributions based on the population, which can often hinder a direct assessment of biological sex as a predictor itself.^28,31^ Anemia and hypertension as risk factors are consistent with the general notion that other systemic comorbidities enhance the susceptibility to viral infection. While anemia may be a marker of general, non-specific disease, hypertension, not generally considered a risk factor for infectious diseases, has emerged as a consistent predictor of SARS-CoV-2 infection and associated complications.^4–7^ The reasons for this are unclear, although angiotensin-converting-enzyme-2 (ACE2)-dependent cellular entry of the virus has been posited as a biologically plausible mechanism, assuming some connection with ACE2-dysregulation that is also associated with hypertension.^32,33^ While optimizing blood pressure control reduces overall morbidity and mortality,^34^ the consequences on incident viral infection have not yet been fully elucidated.

Three at least theoretically readily modifiable exposures, each bolstered by biological plausibility and previous evidence, arose as risk factors for incident viral infection: smoking, household contacts, and the maximum number of recent interactions with other individuals within six feet (about 1·83 meters). The impact of smoking on the risk of SARS-CoV-2 has been difficult to study, as reliance on hospitalization data fails to provide a foundational study base to make comparisons. Smoking may reduce the effectiveness of the immune response and may also upregulate ACE2, rendering individuals more prone to infection.^35^ The observation that sick household contacts predicted incident symptoms may provide evidence that these symptoms were in fact often due to a transmissible disease. For example, although the current analyses excluded those with prevalent symptoms (which reduces the chance symptoms arose from some chronic, ever-present, problem), shared symptoms within a household may have represented some common exposure or predisposition, such as an allergy—however, household symptoms *preceding* participant symptoms arose as a statistically significant predictor of incident symptoms, supporting viral infections as a culprit. Although physical distancing as a method to mitigate spread of infection is supported by the general understanding of the nature of infectious diseases, particularly respiratory viruses, our observation from prospective, repeatedly updated, individual-level data that the number of human to human physical interactions predicted viral symptoms may provide useful evidence in support of physical distancing.

Protective factors included a higher subjective social status, at least weekly exercise, and sanitizing one’s phone. We utilized the MacArthur subjective social status ladder as a validated single-item question to capture socioeconomic status.^19,20^ A higher self-perceived social status may influence viral infection risk in several ways: more education may translate into a better understanding of disease risks and healthy behaviors, and employment among those of a higher socioeconomic status may be more flexible and less often involve high-risk environments. Conversely, stress and the allostatic load related to social determinants of health among those with a lower subjective social status may adversely affect the immune response to infection.^36^ Regular exercise is an established means to improve immune function and the response to viral infection,^37^ now with evidence for beneficial effects specifically in the Covid-19 era. We recognize that sanitizing one’s phone as a protective factor may simply serve as a marker of more fastidious behaviors to minimize risk in general, but the ubiquity and frequent use of the smartphone, likely while shopping, while at work, and while interacting with others, would seem to make it a potentially potent fomite that could result in repeated exposure throughout the day and into the home.

Our study has several important limitations. The outcome of interest was viral symptoms, which relied on self-report. These findings therefore do not directly reflect any particular disease, including infection with SARS-CoV-2. As the study required smartphone use, it is possible our population represents a more technically savvy and perhaps more highly educated and affluent group than the general population. However, this would primarily limit generalizability and should not serve as a threat to internal validity. In addition, the participants were fairly geographically diverse, representing every state in the US and multiple countries. Although less than 80% of the study participants were non-Hispanic white, African American representation was relatively poor. Finally, although the data were collected prospectively and in a time-updated fashion, the study was observational, prone to residual and unmeasured confounding that should temper assumptions of causal effects.

In conclusion, female sex, anemia, hypertension, recent cigarette smoking, living with someone with recent viral symptoms, and the maximum number of recent contacts within six feet (about 1·83 meters) outside the home each predicted a higher risk of developing viral symptoms during the current Covid-19 pandemic. At the same time, a higher subjective social status, regular exercise, and sanitizing one’s phone each predicted a lower risk of developing viral symptoms.

## Data Availability

Interested investigators can submit a proposal to the Eureka Digital Research Platform Executive Committee at UCSF

## Authors’ Contributions

**Conception of the study:** Marcus

**Development of study design:** Marcus, Peyser, Butcher, Olgin, Pletcher, Vittinghoff

**Acquisition of data:** Marcus, Peyser, Joyce, Yang, Tison, Avram, Butcher, Olgin, Pletcher,

**Interpretation of data:** All authors

**Analyses of the data:** Marcus, Avram, Vittinghoff

**Drafting and critical revision of the manuscript:** All authors

**Final approval of the manuscript:** All authors

## Declaration of interests

None of the authors have any pertinent disclosures.

## Acknowledgments

This study was funded by IU2CEB021881-01 and 3U2CEB021881-05S1 from the NIH/ NIBIB to Drs. Marcus, Olgin, and Pletcher.

## Notes

### Competing Interest Statement

The authors have declared no competing interest.

